# Identifying academic success and underperformance: The discriminative power of very short answer questions and multiple-choice questions

**DOI:** 10.64898/2026.04.29.26352108

**Authors:** Elise V. van Wijk, Floris M. van Blankenstein, B.N. Ruijter, J.H.T. Rohling, J. van der Kraan, Friedo W. Dekker, Alexandra M.J. Langers

## Abstract

**Background:** Multiple-choice questions (MCQs) are widely used in medical education, but are criticized for cueing and guessing. Very short answer questions (VSAQs), which require students to generate responses independently, may better assess knowledge. While VSAQs demonstrate higher item discrimination within individual exams, their effectiveness in distinguishing academic performance across multiple assessments remains unclear – representing a key gap in the validation of VSAQs under Messick’s framework, specifically the category of “*relations to other variables*”. This study examines whether VSAQs or MCQs more effectively distinguish students of varying performance levels across multiple summative examinations.

**Methods:** We analyzed retrospective data from six mixed-format examinations with VSAQs and MCQs of three cohorts of first- and second-year medical students. Academic performance was measured using grade point average (GPA) across assessments. Linear regression assessed the relationship of each question format with GPA, while ROC curves and C-statistics evaluated their ability to identify poor and excellent performing students (lowest and highest quintile of GPA).

**Results:** VSAQs showed higher item discrimination (R_ir_-values) than MCQs in all exams. VSAQs also had a stronger positive association with GPA compared to MCQs, and higher C-statistics, indicating superior discriminative ability.

**Conclusion:** VSAQs outperform MCQs in distinguishing academic performance levels across multiple assessments. Their integration into examinations enhances discriminative ability and may facilitate earlier identification of poor and excellent performing students, enabling targeted interventions and support of students.

## Introduction

Assessment of undergraduate medical students predominantly relies on multiple-choice questions (MCQs), especially in the single best answer (SBA) format, due to their high reliability, rapid answering time, and the efficiency of machine marking [1, 2]. However, this format also allows for guessing and cueing (i.e., answering questions by relying on cues from the question stem or answer options rather than on actual content knowledge) and encourages a recognition-based study approach, which may introduce noise into MCQ scores and diminish their ability to accurately reflect students’ true understanding [3]. In contrast, the very short answer question (VSAQ), an open-ended question requiring a concise answer, mitigates these issues by eliminating both cueing and guessing [4]. Consequently, VSAQs tend to exhibit higher item discrimination within individual examinations compared to MCQs [3, 5-7], meaning that they can better differentiate between students with varying levels of performance on a given test. Higher discriminative ability indicates that an item more effectively distinguishes students based on their underlying level of knowledge or competence. Although students need more time to answer VSAQs compared to MCQs, which may reduce the number of questions per exam when testing time is fixed [3], the higher item discrimination of VSAQs implies that fewer questions may be needed per content domain to reliably differentiate between levels of student performance.

Messick conceptualizes validity as a unified construct supported by multiple interrelated resources of evidence, including content, response processes, internal structure, relations to other variables, and consequences of testing. Internal structure refers to the extent to which relationships among test items reflect the intended construct, whereas relations to other variables (external validity) concerns whether test scores relate as expected to other relevant measures, such as academic performance [8]. Establishing both internal and external validity is essential to justify the use of assessment formats in high-stakes educational contexts.

Previous research on VSAQs has primarily focused on internal structure, demonstrating consistently superior item discrimination compared to multiple-choice questions (MCQs) within individual examinations [3, 5–7]. However, higher item discrimination within a single exam does not necessarily translate into a stronger ability to identify students as poor or excellent performers across multiple examinations. This distinction is illustrated by the findings of Eijsvogels *et al*. [9], who showed that extended matching questions (EMQs)– despite superior item discrimination compared to MCQs within examinations [10] – were less effective at identifying poor performing students across multiple assessments. Although EMQs (a form of multiple-choice question in which a single list of answer options is matched to several related clinical scenarios or questions) and VSAQs differ in format, this example underscores that internal structure alone is insufficient to support broader validity claims. Similarly, while VSAQs demonstrate superior item discrimination within individual examinations compared to MCQs [3, 5-7], it remains uncertain whether performance on VSAQs meaningfully relates to broader academic outcomes, which is central to establishing their external validity. Addressing this uncertainty represents a key gap in the validation of VSAQs within Messick’s unified framework.

Beyond evaluating the item discrimination of individual assessment questions, it is therefore important to investigate whether certain question formats are better suited for identifying poor and excellent performing students across multiple examinations. This broader perspective can offer insights into the consistency and robustness of these formats in distinguishing students across different performance levels, and formats with higher discriminative power may facilitate the early identification of underperforming students, thereby enabling timely interventions. In this study, we aim to 1) examine the relationship between question format (VSAQs *versus* MCQs) and academic performance; 2) evaluate the ability of VSAQs and MCQs to identify poor and excellent performing students. To address these aims, we first assess the item discrimination of both question formats within each examination, thereby verifying the assumption that VSAQs have superior discriminative ability within examinations [3, 5-7]. We use the VSAQ- and MCQ-scores from two summative mixed-format examinations administered during the first and second year of an undergraduate medical curriculum. Our analysis includes two student populations: 1) all students who participated in the first-year examination, including those who may later leave the program, and 2) nominal students who participated in both the first- and second-year examinations. The first population offers a broader performance range, particularly among lower performing students, while the second population offers more datapoints (i.e., questions) per student, enhancing the reliability of the analysis.

## Methods

### Setting

This retrospective cohort study was conducted in Leiden University Medical Center (LUMC), the Netherlands. The Dutch medical curriculum comprises a three-year bachelor’s program followed by a three-year master’s program. Although the bachelor courses are primarily assessed with MCQs, other formats such as extended matching, comprehensive integrated puzzle, open essay, and VSAQs are also included in the written assessments. Additionally, students participate in longitudinal training on various CanMEDS competencies [11] beyond the role of Medical Expert, such as communication skills, leadership, health promotion, and collaboration. To assess the discriminative ability of VSAQs and MCQs we analyzed summative examinations of two medical bachelor’s courses: *“Regulation and Metabolism”* (RM), a first-year fundamental course, and *“Diseases of the Abdomen”* (DA), a second-year clinical course. These courses (6 and 7 weeks, respectively) address metabolic and gastrointestinal topics. In our prior study [5], we compared the psychometric properties of MCQs and VSAQs in formative assessments for both courses. Subsequently, the course coordinators added VSAQs to the previously MCQ-only summative exams, resulting in mixed-format examinations. This mixed format has now been used in both courses for three consecutive years (student cohorts 2020-2021; 2021-2022; 2022-2023), resulting in six summative mixed-format examinations available for our analyses. Teaching and exam preparation follows the curriculum program for all cohorts. Students receive scheduled instructional sessions, guided self-study materials, and practice assessments aligned with the exam formats. All students have equivalent access to these learning resources prior to the examination.

All assessments were administered digitally through RemindoToets (Paragin) system [12] and yielded study credits. All teachers involved in the question writing process were required to participate in a course about writing exam questions, where guidelines for writing high quality exam questions were addressed, based on existing literature [13]. Furthermore, all exam questions are reviewed by an Examination Review Board as part of a routine quality assurance system for assessments. VSAQs were marked accordingly to the standard institutional procedure. Predefined correct answers were automatically scored; all other responses were reviewed manually by the examiner. Correct but unanticipated answers (e.g., minor spelling variations) were added to the accepted answer list. Scoring was dichotomous (0/1), with a second examiner consulted when uncertainty arose.

### Participants

The first population, *RM participants*, included all first-year bachelor medical students from the 2020–2021, 2021–2022, and 2022–2023 cohorts who completed the first sitting of the summative RM assessment (*n*=902). The second population, *RM & DA participants*, comprised all second-year bachelor medical students from the same cohorts who completed the first sitting of both the RM and DA summative assessments in consecutive years (*n*=797). Students who attempted less than 75% of all exams taken into consideration for the GPA calculation were excluded from both populations (*n*=51).

### Study design and data collection

We analyzed data from two summative mixed-format examinations from the RM and DA courses, administered between 2021 and 2024. For each assessment, we extracted individual questions, question formats, question scores (coded as 1 for correct and 0 for incorrect), residual item reliability-values (R_ir_) for each question, and student IDs from the Remindo assessment system. The R_ir_-value, representing the correlation between one test item and all other items of the test, measures item discrimination by indicating how well each question correlates with overall student performance on that test (after excluding that specific question) [14]. To assess the comparability of question formats within the examinations, we examined the distribution of each question format across the different themes. The data were accessed between 10/11/2024 and 17/12/2024 for our research. The data were anonymised using ID numbers, ensuring that the authors had no access to information that could identify individual participants.

We measured academic performance with the grade point average (GPA), a widely recognized and standardized indicator of academic success [15, 16]. In our context, GPA is defined as a credit-weighted average of all numeric grades from the first and second year of study. For each student, GPA was calculated based on all courses that included a written examination. In addition to written exams, several courses also comprised other forms of examinations (e.g., oral exams, presentations, or practical assessments) which were scored by a pass or fail, thereby providing a broader representation of academic performance. Courses were weighted according to their European Credit Transfer and Accumulation System (ECTS) credits. The GPA was computed by multiplying each course’s final grade by its corresponding ECTS credits and aggregating these values. All exam grades from the first and second-year courses were retrieved from the university’s administrative system. In our system (used in several European countries), grades range from 1 to 10, with 10 being the highest possible score. A grade of 5.5 or higher is considered a passing grade and is required to earn study credits. Failing grades (defined as <5.5) were included in the GPA calculation to provide a realistic representation of overall academic performance. We evaluated GPA based on two approaches: one using the most recent grade from the last sitting (including retake exams) and another using only the grade from the first sitting. For the first population (*RM participants*) we calculated the GPA based on first-year courses, while for the second population (*RM & DA participants*) the GPA included courses from both the first- and second-year. Based on these GPAs, students were categorized by quintiles into “*poor performing”* (first quintile), *“average performing”* (second through fourth quintile), or*”excellent performing”* (fifth quintile) students.

### Data analysis

Descriptive statistics were calculated and presented for each examination, including total average scores, average VSAQ and MCQ scores, and the distribution of VSAQs and MCQs across the themes. To compare the different question formats, we calculated separate z-scores for VSAQs and MCQs based on their respective absolute scores. For the *RM participants*, z-scores were derived from their VSAQ and MCQ scores on the RM assessment. For the *RM & DA participants*, z-scores were calculated separately for VSAQs and MCQs using absolute scores from both the RM and DA assessments, ensuring that each student had one VSAQ z-score and one MCQ z-score reflecting their performance across both exams. Item discrimination was determined using the mean of the R_ir_-values for each question format [13]. The R_ir_-value for each question was calculated in relation to the entire exam.

Linear regression analysis was performed with GPA as linear outcome variable and the z-scores of the VSAQ and MCQ as covariates, separately for each cohort and across all cohorts. First, univariate regression analyses were conducted to assess the variance explained by each question format individually (reported as adjusted R^2^). Next, both formats were included in a multivariate model to account for shared variance and determine their relative predictive value. Student GPAs from the courses of the first year (*RM participants)* and the first two year (*RM & DA participants)* were used as outcome. Similarly, the same student GPAs were used to create new categorical variables: *“poor performing”* versus *“other”* (average and excellent) and *“excellent performing”* versus *“other”* (average and poor). Receiver Operating Characteristic (ROC) curves were used to evaluate the ability of MCQs and VSAQs to distinguish between poor and non-poor performing students, and also between excellent and non-excellent performing students. The Concordance-statistic (C-statistic or area under the ROC curve) was calculated as a measure of discriminatory power, reflecting how well these formats differentiate between performance levels. Significant differences between the C-statics of VSAQs and MCQs were assessed using paired bootstrapping with 95% confidence intervals. The difference was considered significant when the confidence interval (CI) of the score difference did not contain the zero. Bootstrapping was used to compare the paired C-statistics because it provides robust confidence intervals without relying on strict distributional assumptions. All statistical analyses were performed using R version 4.1.0 (R Foundation for Statistical Computing, Vienna, Austria).

### Ethical approval

This study received ethical approval from the LUMC Educational Research Review Board (OEC/ERRB/20241008/1). Our ethical review board decided that informed consent was not required, as this study did not involve an intervention or experimental manipulation. Rather, VSAQs were introduced as part of an incremental improvement of the existing assessment practice, initially on a small scale to gain experience with this question format. The data analyzed were retrospective and already available from the assessment system as part of the students’ regular curriculum. We collected no additional demographic or personal data, and all data were aggregated and anonymized to ensure that individual students could not be identified.

## Results

### Descriptives and item discrimination

The students included in our study for each mixed-format examination, along with the average total, VSAQ, and MCQ scores are presented in ***Table 1***. In all mixed-format examinations, the average R_ir_-values of the VSAQs were consistently higher compared to the MCQs (***Table 1***). The distribution of question formats across the themes was generally even, with no themes exclusively assessed by a single format (***Supplemental Table 1***).

**Table 1.** The scores and R_ir_-values of VSAQs and MCQs within the mixed-format examinations.

|  | Total examination |  | Very short answer questions |  |  | Multiple-choice questions |  |  |
| --- | --- | --- | --- | --- | --- | --- | --- | --- |
| | Max points | Total score<sup>b</sup><br>mean (SD) | *N* | Score, mean<br>(SD) | $R_{ir}$-values,<br>mean (SD) | *N* | Score, mean<br>(SD) | $R_{ir}$-values,<br>mean (SD) |
| **RM 2021<sup>a</sup>**<br>(*n*=322) | 74.50 | 53.50 (9.00) | 42 | 28.02 (6.26) | 0.30 (0.13) | 32 | 24.53 (3.47) | 0.21 (0.11) |
| **RM 2022**<br>(*n*=295) | 71 | 45.54 (8.76) | 29 | 15.55 (4.62) | 0.28 (0.11) | 36 | 27.31 (4.61) | 0.24 (0.10) |
| **RM 2023**<br>(*n*=285) | 73 | 50.05 (9.61) | 31 | 19.01 (5.34) | 0.32 (0.12) | 38 | 29.33 (4.74) | 0.24 (0.11) |
| **DA 2022**<br>(*n*=282) | 91 | 59.58 (10.35) | 22 | 13.39 (3.41) | 0.30 (0.11) | 48 | 33.34 (4.53) | 0.18 (0.10) |
| **DA 2023**<br>(*n*=255) | 90 | 64.38 (9.48) | 30 | 20.75 (4.14) | 0.26 (0.09) | 40 | 28.50 (3.99) | 0.19 (0.11) |
| **DA 2024**<br>(*n*=260) | 71 | 48.55 (8.18) | 31 | 20.62 (4.18) | 0.27 (0.13) | 20 | 13.66 (2.64) | 0.19 (0.11) |

RM = Regulation and Metabolism; DA = Diseases of the Abdomen; *n* rows = number of students; *N* columns = number of questions.
<sup>a</sup> RM 2021 & DA 2022 = cohort 2020-2021; RM 2022 & DA 2023 = cohort 2021-2022; RM 2023 & DA 2024 = cohort 2022-2023.
<sup>b</sup> Average total score also includes the scores on question formats other than VSAQs and MCQs (comprehensive-integrated-puzzle (CIP) (maximum points: 4), open-ended question without word limitation (maximum points: 8), 6-step pharmacology question (maximum points: 8), hotspot question (maximum points: 1)).

### Relationship between question format and academic performance

We conducted a linear regression analysis to examine the effects of VSAQ and MCQ z-scores on GPA. First, we assessed the variance explained by each question format separately using univariate models (R^2^ values). Among all *RM participants*, the R^2^ values ranged from 0.61 to 0.67 for VSAQ z-score and from 0.44 to 0.58 for MCQ z-score. In all *RM & DA participants*, the R^2^ values ranged from 0.68 to 0.71 for VSAQ z-score and from 0.59 to 0.67 for MCQ z-score.

The multiple regression analyses showed a significant positive association between both VSAQ and MCQ z-scores and GPA (***Table 2***). However, for all *RM participants* (*n*=902), the VSAQ z-score had a stronger association with GPA when using first sitting grades (β = 0.63, t(903) = 21.28, p < .001, 95% CI [0.57, 0.69]) than the MCQ z-score (β = 0.36, t(903) = 12.17, p < .001, 95% CI [0.30, 0.42]), with the model explaining 68% of the variance in GPA (F(2, 899) = 945.79, p < .001, adjusted R^2^ = .68). Similarly, in all *RM & DA participants* (*n*=797), the VSAQ z-score had a stronger positive association (β = 0.53, t(794) = 21.78, p < .001, 95% CI [0.48, 0.57]) compared to the MCQ z-score (β = 0.36, t(794) = 14.84, p < .001, 95% CI [0.31, 0.41]), with the model accounting for 77% of the variance (F(2, 794) = 1303.59, p < .001, adjusted R^2^ = .77). These results were similar when using last sitting grades and for the analyses of the separate cohorts (***Table 2***).

**Table 2.** Linear regression model parameters with grade point average as outcome based on the first and last sitting grades.

| Cohort | First sitting grades |  |  |  |  |  | Last sitting grades |  |  |  |  |  |
| --- | --- | --- | --- | --- | --- | --- | --- | --- | --- | --- | --- | --- |
|  | VSAQ z-score |  |  | MCQ z-score |  |  | VSAQ z-score |  |  | MCQ z-score |  |  |
| | $\beta$ | t | 95% CI | $\beta$ | t | 95% CI | $\beta$ | t | 95% CI | $\beta$ | t | 95% CI |
| *RM participants* |  |  |  |  |  |  |  |  |  |  |  |  |
| 2020-2021<sup>a</sup><br>(n=323) | 0.63 | 14.21 | 0.55–0.72 | 0.25 | 5.64 | 0.16–0.34 | 0.56 | 14.59 | 0.48–0.64 | 0.20 | 5.10 | 0.12–0.27 |
| 2021-2022<br>(n=296) | 0.59 | 11.88 | 0.49–0.69 | 0.46 | 9.17 | 0.36–0.56 | 0.47 | 11.17 | 0.39–0.55 | 0.38 | 8.96 | 0.29–0.46 |
| 2022-2023<br>(n=287) | 0.65 | 12.14 | 0.54–0.75 | 0.38 | 7.20 | 0.28–0.49 | 0.53 | 11.25 | 0.44–0.62 | 0.32 | 6.73 | 0.22–0.41 |
| **Total**<br>**(n=902)** | **0.63** | **21.28** | **0.57–0.69** | **0.36** | **12.17** | **0.30–0.42** | **0.52** | **20.85** | **0.47–0.57** | **0.29** | **11.55** | **0.24–0.34** |
| *RM & DA participants* |  |  |  |  |  |  |  |  |  |  |  |  |
| 2020-2021<br>(n=282) | 0.50 | 11.49 | 0.43–0.58 | 0.34 | 9.06 | 0.27–0.42 | 0.42 | 12.10 | 0.35–0.49 | 0.29 | 8.43 | 0.23–0.36 |
| 2021-2022<br>(n=255) | 0.47 | 13.57 | 0.39–0.55 | 0.39 | 9.52 | 0.31–0.47 | 0.38 | 10.27 | 0.31–0.45 | 0.35 | 9.44 | 0.28–0.42 |
| 2022-2023<br>(n=260) | 0.59 | 14.21 | 0.51–0.68 | 0.35 | 7.90 | 0.26–0.43 | 0.52 | 13.48 | 0.44–0.59 | 0.28 | 7.44 | 0.21–0.36 |
| **Total**<br>**(n=797)** | **0.53** | **21.78** | **0.48–0.57** | **0.36** | **14.84** | **0.31–0.41** | **0.44** | **20.38** | **0.40–0.48** | **0.31** | **14.14** | **0.26–0.35** |

RM=Regulation and Metabolism; DA = Diseases of the Abdomen; VSAQ=very short answer question; MCQ=multiple-choice question; CI=confidence interval. SE = standard error. All p-values <0.001. <sup>a</sup> Cohort 2020-2021 = RM 2021 & DA 2022; Cohort 2021-2022 = RM 2022 & DA 2023; Cohort 2022-2023 = RM 2023 & DA 2024.

### Identification of poor and excellent performing students

We calculated the C-statistic to assess the ability of VSAQ and MCQ scores to identify poor performing (lowest GPA quintile) and excellent performing (highest GPA quintile) students. In all *RM participants* (*n=*902), VSAQ z-scores showed higher C-statistics (i.e., greater discriminative ability) for both poor and excellent performing students compared to MCQ z-scores (***Figure 1A***). However, for poor performing students using first sitting grades, the difference between the C-statistics of VSAQ and MCQ was not significant. In all *RM & DA participants* (*n*=797), the C-statistic of the VSAQ z-score remained significantly higher than that of the MCQ z-score for poor performing students using first sitting grades. For excellent performing students, there was no significant difference in discriminative ability (***Figure 1B***). The findings across separate cohort analyses followed the same pattern; either the C-statistic for the VSAQ z-score was significantly higher than that of the MCQ z-score, or there was no significant difference between them (***Supplemental Figure 1*** & ***2***).

**Figure 1.**
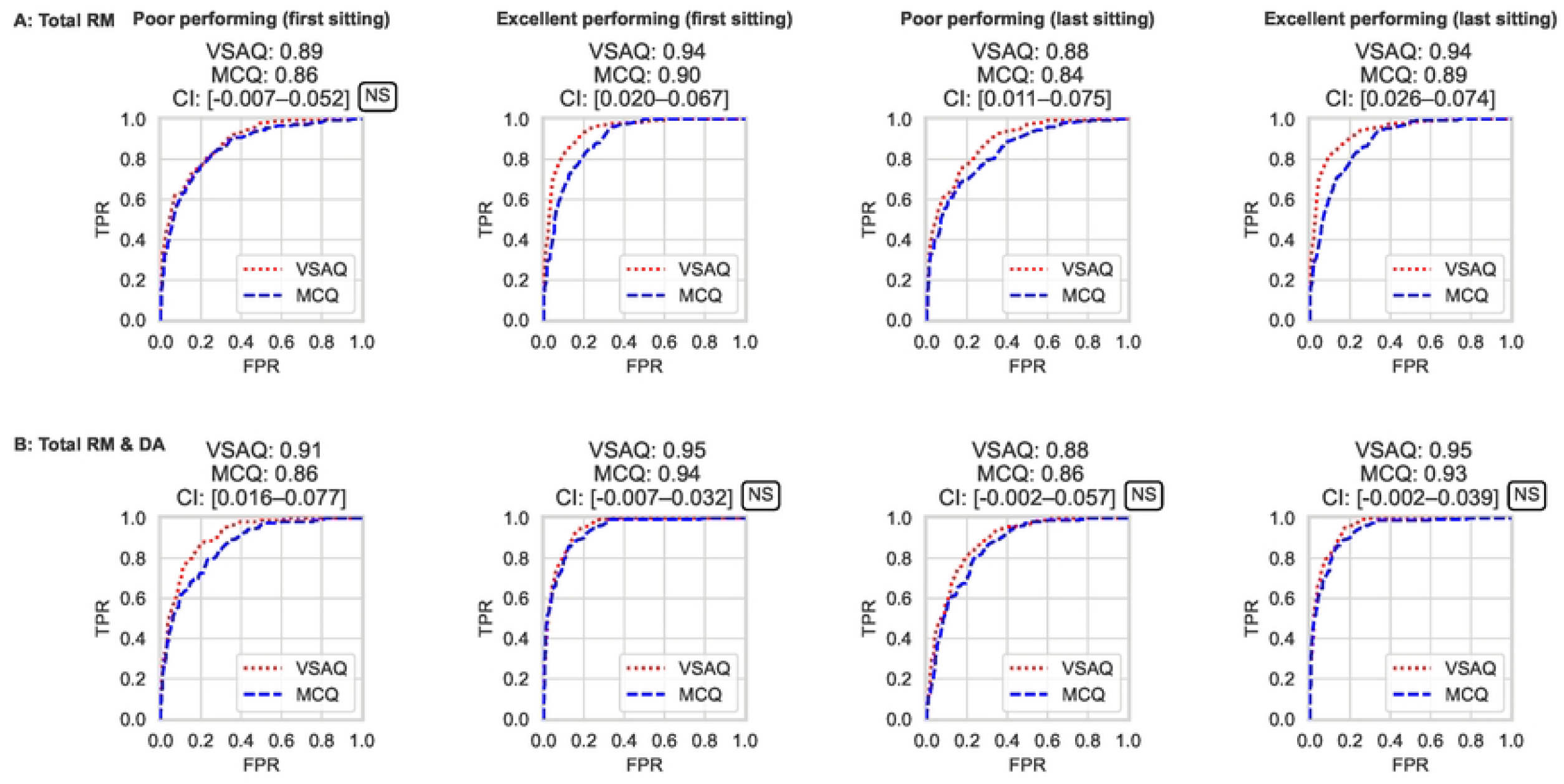
Receiver Operating Characteristic (ROC) curves for both poor and excellent performing students using the first sitting or last sitting grades from **A)** the total RM participants, and **B)** the total RM & DA participants. Red = VSAQs; Blue = MCQs; CI = 95% confidence interval; TPR = true positive rate; FPR = false positive rate. NS = non-significant. C-statistics are shown above each graph, which were calculated from the area under the ROC curve.

## Discussion

In this study, we examined the relationship between question format (VSAQs *versus* MCQs) and academic performance based on GPA, and evaluated the ability of both formats to distinguish poor-performing students from all other students, and excellent-performing students from all other students. Our findings reveal that while both question formats are suitable for distinguishing students across performance levels, VSAQs consistently demonstrate superior effectiveness compared to MCQs in both the linear regression analysis and ROC curves. Notably, the difference in discriminative ability between VSAQs and MCQs was more pronounced among first-year students (*RM participants*) compared to the combined group of first- and second-year students (*RM & DA participants*). A possible explanation for this observation is the exclusion of mostly poor performing students who did not nominally progress to the second year, resulting in a more homogeneous group of students with less variability in academic performance. Consistent with previous research [3, 5-7], VSAQs also demonstrated higher item discrimination than MCQs within the individual examinations. Our findings complement those of Eijsvogels *et al*. [9] by examining a different question format – VSAQs – that shares certain psychometric features with EMQs, but may function differently in practice. The observed differences in outcomes likely reflect format-specific characteristics. Although EMQs reduce reliance on guessing compared to traditional MCQs, they still provide a list of plausible options that could lead to guessing or cueing. In contrast, VSAQs require students to generate concise responses without external cues, inherently minimizing the likelihood of guessing and offering a more direct measure of their knowledge. Furthermore, Eijsvogels *et al*. [9] penalized incorrect answers on MCQs, which may have enhanced the discriminative performance of this question format. By normalizing performance scores within mixed-format examinations, we ensured a fair comparison, eliminating potential biases from variations in exam quality or instructional approach. Additionally, our analysis included multiple examinations and a larger sample size, enhancing the robustness of our findings.

The GPA used in this study to assess academic performance is primarily based on assessments that predominantly consist of MCQs, which could bias the results toward MCQs due to their alignment with the format used to calculate GPA. However, despite this potential bias, our findings indicate a clear advantage for VSAQs over MCQs in distinguishing poor and excellent performing students. The superior performance of VSAQs can be attributed to their format, which requires students to generate responses independently, eliminating opportunities for guessing and relying instead on their actual understanding [17-20]. This characteristic makes VSAQs a more accurate reflection of student’s knowledge level. While guessing may inflate MCQ scores for poor performing students in a single examination, this effect will diminish when performance is averaged across multiple exams, thereby reducing noise introduced by guessing.

### Strengths and limitations

This study is the first to evaluate the discriminative ability of VSAQs across multiple assessments using GPA as a measure of academic performance (evidence of relations to other variables), rather than focusing exclusively on item discrimination with individual exams (evidence of internal structure). By utilizing real-world summative assessment data from three distinct student cohorts, we minimized the biases associated with voluntary participation and ensured a robust, and representative sample size. This also allowed us to aggregate results and average out variance across examinations and populations, thereby enhancing the reliability of our findings. The inclusion of both first-year students with greater variation in academic performance, and nominal second-year students allowed for a more comprehensive analysis of performance across different populations, while mitigating selection bias. Moreover, mixed-format examinations enabled within-subjects comparisons of VSAQs and MCQs, under comparable instructional and assessment conditions. Standardizing scores further reduced variability and ensured reliable comparisons. However, this study also has limitations. While GPA is an objective and widely used measure of academic achievement, it simplifies complex learning outcomes and may overlook critical thinking and skill development [15]. Additionally, the variability in content and distribution between VSAQs and MCQs within real-world examinations could introduce bias. However, we mitigated this by aggregating data from multiple examinations and cohorts, standardizing scores, and conducting analyses across two student populations.

### Implications for practice and future research

Our findings indicate that VSAQs have a higher discriminative ability than MCQs, effectively distinguishing students both within individual examinations and across multiple examinations based on GPA. This suggests that VSAQs provide a more robust and valid question format for evaluating academic performance, supporting their implementation to enhance the discriminatory power and reliability of assessments. Incorporation of VSAQs into assessments might also enhance the ability to identify students in need of early interventions and support, while also recognizing those who excel. Moreover, if our findings are confirmed by future studies, VSAQs could be considered as a potential tool in medical school selection by assessing study success potential while simultaneously providing an authentic preparation for the curriculum. Teachers could leverage VSAQs to implement tailored strategies for improving learning outcomes and to identify exceptional students for advanced opportunities. Future studies could expand academic performance measures beyond GPA, incorporating assessments of skill development and work-place learning. These additional metrics would provide a more holistic evaluation of the effectiveness of VSAQs and MCQs across different educational contexts. Additionally, exploring whether VSAQs are associated with long-term academic and professional success, including performance in real-world clinical settings, would provide deeper insights into their utility.

### Conclusion

This study suggests that VSAQs may be more effective than MCQs in distinguishing undergraduate medical students across different academic performance levels. Our findings indicate that VSAQ scores are associated with broader academic performance beyond the exam itself, contributing to the *“relations to other variables”* dimension of Messick’s unified validity framework. Integrating VSAQs into assessments may enhance the discriminative power and robustness of assessments and could support early identification of both poor and excellent performing students, potentially enabling more targeted educational support. Future research is needed to explore the generalizability of these findings across diverse educational settings and to evaluate the extent to which VSAQs may predict long-term academic achievements.

## Data Availability

The anonymized data and accompanying code used for the results in this study are freely available from the GitHub repository (https://github.com/elisevanwijk/van_wijk_2026_plos_one).

## Supporting information

**Supplemental Table 1.** Distribution of VSAQs and MCQs across the themes within the examinations

**Supplemental Figure 1**. Receiver Operating Characteristic (ROC) curves for both poor and excellent performing students using the first sitting or last sitting grades from the RM participants cohort 2020-2021 (RM 2021), cohort 2021-2022 (RM 2022), and cohort 2022-2023 (RM 2023).

**Supplemental Figure 2**. Receiver Operating Characteristic (ROC) curves for both poor and excellent performing students using the first sitting or last sitting grades from the RM & DA participants cohort 2020-2021 (RM 2021 & DA 2022), cohort 2021-2022 (RM 2022 & DA 2023), and cohort 2022-2023 (RM 2023 & DA 2024).

## Acknowledgements

The authors wish to thank all study coordinators and teachers for their critical appraisal of the research protocol and valuable contribution to the manuscript.

